# On Reliability of the COVID-19 Forecasts

**DOI:** 10.1101/2020.06.01.20118844

**Authors:** Hemanta Kumar Baruah

**Affiliations:** Department of Mathematics, The Assam Royal Global University, Guwahati, Assam - 781035, INDIA

**Keywords:** Corona virus disease, mathematical modeling, extrapolation, forecasting, 33F05, 93A30

## Abstract

In this expository article, we are aiming to show with an example that even short term forecasts regarding the COVID-19 spread pattern may sometimes not be very reliable. We have studied data published by Worldometers.info to get numerically an approximate formula of the spread pattern for a short period. We have observed that in the United States of America, there was a nearly exponential spread pattern for a very short period from May 3 to May 8, 2020. From May 9 to May 13, the nearly exponential character of the spread was found to be absent. Hence it can be concluded that the COVID-19 spread pattern, even after more than four months from the start of the outbreak, is not quite predictable. Therefore even short term forecasts regarding the spread may not be very reliable. We have found that forecasts using the assumption of an exponential pattern of spread may actually lead to overestimation.

## 1. Introduction

After the COVID-19 outbreak, many authors have studied the spread pattern of the virus. Various mathematical techniques such as time series analysis and simulation have been used for forecasting the spread pattern of the virus. In the available literature, data from the initial stage of the outbreak have been analyzed in detail, and predictions regarding the spread have been made for different countries.

By now the virus has made inroads into almost all countries. However, while in some countries the pandemic is subsiding, in some others it is still in the initial stage. Accordingly, if an attempt is made to make a forecast regarding the spread pattern considering the world as a whole, a situation may occur that in a country that had been contributing highly towards the spread may suddenly start to show a different trend. That might affect the forecasts made keeping in view the entire world. The objective of this work is to show with an example using numerical explanations that even short term forecasts regarding the COOVID-19 matters may not actually be very reliable.

Various mathematical models for studying the spread pattern of epidemics are available in the literature. Such models, framed using certain assumptions, are explained with the help of differential equations. Meyers [1] used the Susceptible-Infected-Recovered (SIR) model to study the forecasts of small outbreaks. Lutz *et. al*. [2] concluded that the use of infectious disease forecasts for decision making may be challenging because such forecasts are neither standardized nor validated and may be difficult to communicate to people. These two works on infectious disease forecasting were published much before the COVID-19 outbreak.

We shall now cite some of the works on COVID-19 spread patterns published very recently. The work of Wu *et. al* [3] published on 31 January, 2020, was perhaps one of the earliest works done on the novel corona virus spread. They used the susceptible-exposed-infectious-recovered (SEIR) model to make a simulation study of the spread. The work of Al Hassan *et. al*. [4], published on 17 February, 2020, was another work done on COVID-19 spread in its initial stage. Using trend analysis, they had observed that cases were increasing at a reduced rate, whereas death had been increasing continuously. Kucharski *et. al*. [5] studied the dynamics of transmission of COVID-19 in the very early stage, in February, 2020. They performed Monte Carlo simulations for mathematical modeling. Kuniya [6] used the SEIR model for prediction of the peak of the corona virus disease, and concluded that intervention over a relatively long period would be needed to reduce the pandemic size effectively. Binti Hamzah *et. al*. [7] used the SEIR predictive modeling using the data of China and outside, and concluded that further studies need to be done to help to contain the outbreak as early as possible. Regarding estimation of the spread pattern of the pandemic, Gondauri *et. al*. [8] concluded that it is difficult to predict about the spread as on March 30. A clearly observable pattern had not anyway emerged by that time. Anastassopoulou *et. al*. [9] used the Susceptible-Infectious-Recovered-Dead (SIRD) model to do a simulation study towards predicting the situation in Hubei by the end of February. Li *et. al*. [10] used the Gaussian distribution to construct a model of corona virus transmission. They used data available at the initial stage of the outbreak to study the Hubei related matters only. Petropoulos and Makridakis [11] have made five rounds of forecasts covering the period from February 2, 2020, to March 21, 2020, and in the analysis and forecasting for the fourth round, for data from March 2, 2020, to March 11, it was mentioned that they were under-forecasting the real situation, and that was due to the exponential increase of the confirmed cases in Europe, Iran and the United States. They have made a forecast of continuous increase in the COVID-19 cases. Perc *et. al*. [12] have concluded that the pandemic was growing exponentially in various countries around the world, including the United States. Ciufolini and Paolozzi [13] studied the data from China and Italy available at the end of March, 2020, and concluded that the Gaussian distribution can be used for prediction of the number of cases. Chintalapudi *et. al*. [14] studied the outbreak pattern using the auto-regressive integrated moving average method of time series analysis. Kim *et. al*. [15] used the SEIR model to study the COVID-19 spread in Korea and estimated the total number of cases in Korea by the middle of June.

It may be noted the data analyzed in the works cited above were of the initial period of the outbreak. These works were basically aimed at forecasting the spread pattern in certain countries. Slowly and steadily, the virus entered into almost all countries of the world. So by now, many patterns got imbedded in the data of the total number of cases in the world. By the time in some countries the outbreak had started to take a predictable shape, newer entries were in the initial stage of the outbreak. From one country to another, the spread patterns are currently different. Therefore a clear prediction for the world as a whole may not be possible simply because the countries are in different stages of the pandemic. Further, application of the SEIR type of models is proper only in the cases in which we consider a particular country with specific required facilities to cope up with the outbreak. When we consider the totality of the cases around the world, such facilities are obviously different in different countries and therefore forecasting regarding the total number of cases considering the world as a whole using the SEIR type of models is not actually feasible.

## 2. Methodology

In what follows, we are now going to discuss why we are raising the question of reliability of the forecasts regarding the COVID-19 spread. We shall use data published by Worldometers.info [16] regarding the total number of cases that includes active cases, recovered cases and deaths. To discuss our standpoint we shall consider the total number of cases in the United States. From the Worldometers.info data, it can be seen that there was a nonlinear increasing trend in every country including the United States right from the beginning and very soon the pattern became highly nonlinear.

Petropoulos and Makridakis [11] have mentioned that in Europe, Iran and USA, the spread was exponential. Perc *et. al*. [12] also have given a similar comment that the pandemic was growing exponentially in countries including USA. Both of these two works were based on the data of the initial period of the pandemic. We are interested to study how far this claim of exponential increase is acceptable even during this current period.

Let us assume that the total number *N* of cases is following an exponential pattern on any given day *t*. If *N* is a function of *t* following

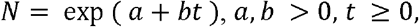

with *a* and *b* constants, then *Y* = *log_e_ N* is linear in *t*. Now as (*a*+*bt*) is a first degree expression, the first order differences Δ*Y* of *Y* = (*a* + *bt*) would be very nearly constant for equidistant values of *t*.Therefore if we see that the first order differences Δ*Y* of Δ*Y* of observed values of *Y* for equidistant *t* are approximately constant, then we may say that *N* is nearly exponentially increasing. It is obvious that Δ*Y* can be only approximately constant because the values of *Y* would include random errors. Further, approximate constancy of Δ*Y* would be there only if *Y* is approximately linear in *t*.

At this point, we would like to mention one particular point. There is a time series data set available regarding the COVID-19 spread. We would look into the matters from May 8 downwards to find out approximately from which date the spread pattern in USA was very nearly exponential. It is certain that the exponential character could not have been there right from the beginning. Further, the parameter *b* in *N* = exp(*a* + *bt*) cannot really be a constant anyway, it has to be time dependent, linearly possibly. Therefore if we consider the values of *N* from a recent date downwards, we would be able to find how *b* is behaving in a particular small period of time. Our method of analysis would be to be look into the time series from May 8 downwards, and we would like to see whether there is any nearly exponential pattern in the data.

## 3. Results and Discussions

To discuss about reliability of the forecasts, we would now like to show the COVID-19 spread data of USA for a short period. In the two tables below, Tables-1 and 2, we would show the values of *N*, *Y* = *log_e_N* and Δ*Y* for the United States in two specific periods. For USA, our data source Worldometers.info [16] has depicted the values in a graph wherefrom the numbers can be found immediately after clicking on the graph. At this point we would like to mention that we are not really going to use some standard statistical procedure for forecasting the total number of cases, we are interested only to show numerically that the forecasts may actually be very high overestimations of the reality.

**Table-1:** Values of Δ*Y* for USA from May 3 to May 8.

| Date | $N$ | $Y$ | $\Delta Y$ |
| --- | --- | --- | --- |
| May 8 | 1321785 | 14.09449 | 0.02230 |
| May 7 | 1292623 | 14.07218 | 0.02311 |
| May 6 | 1263092 | 14.04907 | 0.02036 |
| May 5 | 1237633 | 14.02871 | 0.02024 |
| May 4 | 1212835 | 14.00847 | 0.02058 |
| May 3 | 1188122 | 13.98788 | 0.02331 |

**Table-2:**
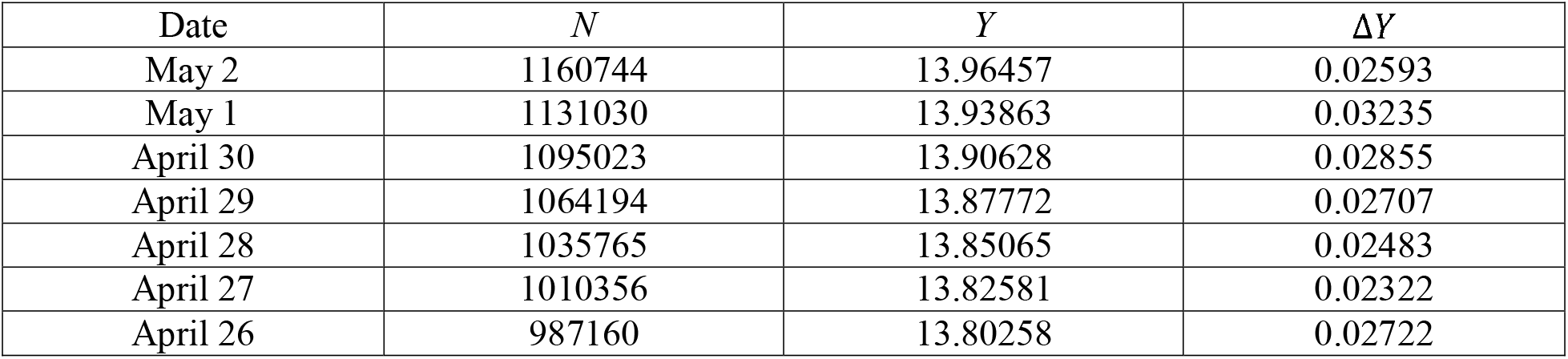
Values of Δ*Y* for USA from April 26 to May 2

| Date | $N$ | $Y$ | $\Delta Y$ |
| --- | --- | --- | --- |
| May 2 | 1160744 | 13.96457 | 0.02593 |
| May 1 | 1131030 | 13.93863 | 0.03235 |
| April 30 | 1095023 | 13.90628 | 0.02855 |
| April 29 | 1064194 | 13.87772 | 0.02707 |
| April 28 | 1035765 | 13.85065 | 0.02483 |
| April 27 | 1010356 | 13.82581 | 0.02322 |
| April 26 | 987160 | 13.80258 | 0.02722 |

It may be observed that in Table-1, the values of Δ*Y* are very nearly constant, which means that *Y* for USA from May 3 to May 8 were very nearly linear, and therefore the total number of cases *N* is approximately of the exponential type with

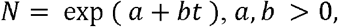

from May 3 to May 8. However, in Table-2 we find that the values of Δ*Y* cannot be said to have even approximate constancy, and therefore during this period the pattern was far from being exponential although the values of *N* were very highly nonlinear in this table.

Now this leads us to conclude that the data regarding the total number of COVID-19 cases in USA were not of the exponential type at the end of March as Petropoulos and Makridakis [11] and Perc *et. al*. [12] have concluded. As for unreliability of the forecasts, we shall first present the values of *N* for the period May 9 to May 13. We would like to show that the spread pattern in USA abruptly changed after May 8. From May 3 to May 8, as shown in Table-1, Δ*Y* was nearly linear. After May 8, it can be observed that *N* was indeed increasing but it was not increasing exponentially like it was in the period May 3 to May 8. In fact the rate of increase of *N* suddenly started to diminish observably after May 8.

Taking May 3 as the base and with Δ*Y* = 0.02331,the value of Δ*Y* for May 3, we have checked that the interpolated values for May 4 to May 8 are very close to the observed values. This shows that from May 3 to May 8 the nearly exponential model does work well.

Let us now hypothesize that the COVID-19 spread in USA is nearly exponential after May 8. Now if we take May 8 as the base and calculate the values of *N*, we shall see that forecasts from May 9 to May 13 would be as shown in Table-3. For the purpose of extrapolation, we shall proceed as follows: The value of *Y* on May 8 was 14.09449. We shall assume that Δ*Y* = 0.02165, the mean of the values ofΔ*Y* in Table-1. Therefore the calculated value of *N* for May 9 would be

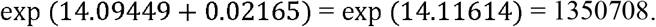

The process can be continued from May 10 to May 13 to get the other values.

**Table-3:** Calculated and Observed Values of *N* in USA, May 9 to May 13

| Date | Calculated $Y$ | Calculated $N$ | Observed $N$ |
| --- | --- | --- | --- |
| May 9 | 14.11614 | 1350708 | 1347309 |
| May 10 | 14.13779 | 1380270 | 1367638 |
| May 11 | 14.15944 | 1410479 | 1385834 |
| May 12 | 14.18109 | 1441348 | 1408636 |
| May 13 | 14.20274 | 1472894 | 1430347 |

As can be seen from Table-3, the calculated values with May 8 as the base have been found to overestimate the total number of cases. This happened because the spread of COVID-19 in USA has started to be slower after May 8.

In fact, if we go for a test of goodness of fit using the *χ*^2^ test of significance, we can see that the tabulated values of *χ*^2^ for 4 degrees of freedom at 5% and 1% levels of significance are 9.488 and 13.277 respectively, whereas our calculated value of *χ*^2^ is 2526.229. Hence we can conclude that we have to reject our hypothesis that the spread pattern was nearly exponential in USA after May 8. Therefore estimates that had been based on an assumption that the total number of cases has been growing exponentially are not quite reliable.

Indeed, from the Worldometers.info data, it can be observed that not just in USA, in countries such as Spain and Italy also the spread had changed at some points in the same pattern. It may be that due to precautions taken and as a result of lockdown, the spread rate diminished slowly in these countries. These countries had been contributing towards the total number of cases daily in large numbers. As such even if the spread pattern in the world as a whole might at the moment look nearly exponential, a prediction based on that even for a short term may actually lead to an overestimation. What we mean is that if we forecast the total number of cases in the world assuming that the spread pattern is nearly exponential taking May 8 as the base, we would end up overestimating the situation because there was a decline in the total number of cases in USA after May 8 and at that time the highest daily increase in the total number was from USA.

## 4. Conclusions

Predictions regarding COVID-19 spread may not be very perfect. In fact, the predictions that are based on an assumption that the spread is exponential may actually end up overestimating the situation. Therefore even short term forecasts regarding the spread considering the world as a whole may not be very reliable. This is so because the countries that had been contributing in very large numbers towards increase in the total number of COVID-19 cases has now started to be in a stage in which the rate of growth actually has a decreasing trend already.

## Data Availability

The data have been taken from Worldometers.info.

https://www.worldometers.info/coronavirus/coronavirus-cases/

## Notes

### Competing Interest Statement

The authors have declared no competing interest.

### Clinical Trial

This work is not on clinical trials.

### Funding Statement

This work is not supported by any funding agency.

### Author Declarations

This work does not need any approval of the IRB.

